# Axitinib in Progressive Metastatic Pheochromocytoma and Paraganglioma: Results of a Phase II Study

**DOI:** 10.64898/2026.09.28.26364204

**Authors:** Jaydira Del Rivero, Andrea B. Apolo, Maureen Edgerly, W Douglas Figg, James Lee, John Ausiello, Karel Pacak, Susan E. Bates, Tito Fojo

## Abstract

**Purpose:** Metastatic pheochromocytoma and paraganglioma (PPGL) are rare neuroendocrine malignancies with limited systemic treatment options. Although vascular endothelial growth factor (VEGF) pathway inhibition has demonstrated activity in metastatic PPGL, prospective data evaluating selective VEGF receptor inhibition are limited. Axitinib is a potent, selective inhibitor of VEGFR-1, -2, and -3. We evaluated the efficacy and safety of axitinib in patients with progressive metastatic PPGL.

**Methods:** Patients with progressive metastatic or unresectable PPGL were enrolled in a prospective, open-label, single-arm, phase II study (NCT03839498). Eligible patients had measurable disease by RECIST version 1.1 and no prior treatment with a tyrosine kinase inhibitor. Axitinib was administered orally at an initial dose of 5 mg twice daily with protocol-defined dose modifications based on tolerability. The primary end points were objective response rate (ORR) and progression-free survival (PFS). Adverse events were graded according to the National Cancer Institute Common Terminology Criteria for Adverse Events (CTCAE).

**Results:** Nineteen patients were enrolled, and 17 received study treatment and were evaluable for efficacy and safety. Six patients achieved a confirmed partial response, with an ORR of 35.3%. Median progression-free survival was 7.9 months (95% CI, 6.1-16.8 months), median duration of response was 7.4 months, median duration of treatment was 9.5 months, and median overall survival was 29 months. Hypertension, the most common treatment-related adverse event, occurred in 79% of patients, was effectively managed with antihypertensive therapy and dose modifications. Other common treatment-related adverse events included fatigue, diarrhea, oral mucositis, and palmar-plantar erythrodysesthesia. No patient permanently discontinued treatment because of treatment-related toxicity.

**Conclusions:** Axitinib demonstrated clinically meaningful antitumor activity in patients with progressive metastatic PPGL with a safety profile consistent with VEGF pathway inhibition and manageable with supportive care and dose modification. The observed activity was comparable to that reported with other VEGF-targeted therapies, including sunitinib. These findings support axitinib as a potential treatment option for patients with metastatic PPGL and warrant further evaluation.

## Translational Relevance

Metastatic pheochromocytoma and paraganglioma (PPGL) are rare neuroendocrine malignancies with limited systemic treatment options and few prospective clinical trials. Aberrant angiogenesis driven by vascular endothelial growth factor (VEGF) signaling is a characteristic feature of PPGL considered by many a hallmark of these tumors, particularly in pseudohypoxia-associated molecular subtypes, including SDHx-mutant disease, providing a biological rationale for VEGF receptor inhibition. [1, 2] Although the randomized phase II FIRSTMAPPP trial established sunitinib as an active treatment option, and belzutifan has recently become the first FDA-approved oral therapy for PPGL, additional effective therapies with distinct kinase selectivity and toxicity profiles remain needed.[3-5]

This prospective phase II study evaluated axitinib, a potent and selective inhibitor of VEGFR-1, VEGFR-2, and VEGFR-3 in patients with progressive metastatic PPGL. Axitinib demonstrated clinically meaningful antitumor activity and the study provides detailed characterization and management of treatment-related toxicities, including the management of hypertension and other VEGF pathway-associated adverse events in a population with underlying catecholamine excess. The findings demonstrate that these toxicities can generally be managed with supportive care, antihypertensive therapy, and protocol-directed dose modification, with no permanent treatment discontinuation because of treatment-related toxicity.

These data expand the prospective clinical evidence supporting VEGF-directed therapy in metastatic PPGL, identify axitinib as a potential therapeutic option, and provide practical information regarding toxicity management that may inform the clinical management of this rare endocrine malignancy.

## INTRODUCTION

Pheochromocytomas and paragangliomas (PPGLs) are rare neuroendocrine neoplasms arising from chromaffin cells of the adrenal medulla and extra-adrenal paraganglia, respectively. Although most PPGLs follow an indolent clinical course, approximately 15%-30% ultimately develop metastatic disease, which remains the principal cause of disease-related mortality. [6, 7] Unlike many solid tumors, metastatic PPGL is defined by the presence of tumor deposits at sites where normal chromaffin tissue is absent rather than by histologic features alone. Patients with metastatic PPGL may experience substantial morbidity related to both tumor progression and catecholamine excess, when present, resulting in hypertension, tachyarrhythmias, cardiovascular complications, pain, and reduced quality of life. [8, 9] Consequently, treatment goals include durable tumor control while minimizing treatment-related toxicity and maintaining clinical stability.

Historically, systemic treatment options for metastatic PPGL have been limited. Combination chemotherapy with cyclophosphamide, vincristine, and dacarbazine (CVD) demonstrated activity with modest response durability. [10, 11] Radiopharmaceutical therapies, including high-specific-activity ^131^I-metaiodobenzylguanidine (^131^I-MIBG) and peptide receptor radionuclide therapy (PRRT), have expanded therapeutic options for appropriately selected patients; however, not all tumors demonstrate sufficient tracer uptake, and durable responses remain uncommon. [12] More recently, the HIF-2α inhibitor belzutifan has demonstrated durable antitumor activity in the LITESPARK-015 trial and received FDA approval in May 2025 for locally advanced, unresectable, or metastatic PPGL, becoming the first oral therapy approved for this indication. [13]

The molecular pathogenesis of PPGL has become increasingly well characterized. Germline pathogenic variants are identified in approximately 40% of patients, making PPGL one of the most heritable human neoplasms. [14, 15] Tumors harboring pathogenic alterations in genes involved in cellular oxygen sensing and mitochondrial metabolism, including SDHA, SDHB, SDHC, SDHD, FH, VHL, and EPAS1, demonstrate activation of pseudohypoxia signaling pathways characterized by stabilization of hypoxia-inducible factors (HIFs) and increased expression of vascular endothelial growth factor (VEGF). [13, 16, 17] This angiogenic phenotype provides a biologic rationale for therapeutic inhibition of VEGF signaling. SDHX-associated tumors are of particular interest because of their association with pseudohypoxia signaling and angiogenesis, providing further rationale for evaluation of VEGF-targeted therapies. [18, 19]

Several prospective and retrospective studies have examined the clinical activity of multitargeted TKIs, including sunitinib, cabozantinib, pazopanib, and lenvatinib.[4, 20] Among these, the randomized, placebo-controlled FIRSTMAPPP trial established sunitinib as the first agent to demonstrate a significant improvement in progression-free survival in patients with progressive metastatic PPGL, providing prospective evidence supporting VEGF-targeted therapy in this disease. [1], [18] More recently, a meta-analysis of TKIs in metastatic PPGL reported a pooled partial response rate of 32% and a pooled median PFS of 8.9 months. [12] However, treatment-related toxicities associated with multitargeted kinase inhibition frequently necessitate dose reductions, treatment interruptions, or discontinuation, emphasizing the need for additional agents with greater kinase selectivity and potentially improved tolerability. [2]

Axitinib (AG-013736) is an orally administered second-generation tyrosine kinase inhibitor with high potency and selectivity for VEGF receptors 1-3. [21] Compared with earlier multitargeted TKIs, axitinib has relatively limited activity against non-VEGF kinases, providing a rationale for investigating whether more selective VEGF receptor inhibition can retain antitumor activity with a manageable toxicity profile. [22, 23]. Given the central role of VEGF signaling in metastatic PPGL, we conducted a prospective, open-label, single-arm phase II study to evaluate the efficacy and safety of axitinib in patients with progressive metastatic PPGL. We hypothesized that selective VEGF receptor inhibition with axitinib would provide clinically meaningful antitumor activity with a manageable safety profile.

## MATERIALS AND METHODS

### Study Design

This study was a prospective, open-label, single-arm, phase II clinical trial evaluating axitinib in patients with progressive metastatic PPGL (ClinicalTrials.gov identifier NCT03839498). The primary end point was objective response rate (ORR). Secondary end points included progression-free survival (PFS), duration of response, overall survival, treatment duration, and safety.

The study protocol was approved by the Institutional Review Boards of the National Cancer Institute and Columbia University and was conducted in accordance with the principles of the Declaration of Helsinki, the International Council for Harmonisation Good Clinical Practice guidelines, and all applicable federal regulations. Written informed consent was obtained from all participants before enrollment.

### Patient Population

Adults (≥18 years) with histologically confirmed pheochromocytoma or paraganglioma and metastatic, unresectable, or locally advanced disease were eligible. Eligibility required radiographically documented disease progression before study entry, at least one measurable lesion according to Response Evaluation Criteria in Solid Tumors (RECIST) version 1.1, an Eastern Cooperative Oncology Group (ECOG) performance status of 0-2, and adequate organ function. Biochemical confirmation of catecholamine-producing disease was required when clinically applicable. Patients who had previously received a VEGF receptor tyrosine kinase inhibitor were excluded, as were those with uncontrolled cardiovascular disease, active infection, or clinically significant bleeding diathesis. Patients receiving at least one dose of study treatment were included in the safety analysis. Patients who completed at least one post-baseline radiographic assessment were included in the efficacy analysis.

### Treatment Administration and Dose Modification

Axitinib was administered orally at a starting dose of 5 mg twice daily on a continuous dosing schedule in 28-day cycles. Treatment continued until radiographic disease progression, unacceptable toxicity, withdrawal of consent, investigator decision, sponsor-initiated study termination, or study closure. Patients who tolerated the starting dose without clinically significant toxicity for at least four consecutive weeks were eligible for stepwise dose escalation to 7 mg and then 10 mg twice daily. Conversely, the dose could be reduced to 3 mg, and subsequently2 mg, twice daily for treatment-related toxicity. Temporary treatment holds were permitted until toxicity resolved to an acceptable level, after which axitinib could be resumed at the same or a reduced dose at the discretion of the treating investigator. Concomitant supportive care, including antihypertensive therapy, thyroid hormone replacement, electrolyte repletion, analgesics, and antiemetics, was provided per institutional standards.

### Assessments

At baseline, all patients underwent a comprehensive clinical and laboratory evaluation. During treatment, clinical and laboratory evaluations were performed every four weeks. Blood pressure was recorded at each visit and monitored more frequently when clinically warranted. Tumor response was assessed every 12 weeks (±2 weeks) by computed tomography or magnetic resonance imaging and evaluated by study investigators according to RECIST version 1.1.

### End Points

The primary end point was objective response rate (ORR), defined as the proportion of patients achieving a confirmed complete or partial response per RECIST 1.1. Secondary endpoints included progression-free survival (PFS), defined as the time from the first dose of axitinib to radiographic progression or death from any cause, whichever occurred first; duration of response (DOR), disease control rate (DCR), defined as the proportion of patients achieving complete response, partial response, or stable disease lasting at least 12 weeks; overall survival (OS), defined as the time from first dose to death from any cause; and duration of treatment.

### Safety Evaluation

Adverse events were recorded continuously from the first dose through 30 days after the last dose and graded according to the National Cancer Institute Common Terminology Criteria for Adverse Events (CTCAE) version IV. The treating investigator assigned each event an attribution score on a five-point scale: unrelated (1), unlikely (2), possible (3), probable (4), or definite (5). For this analysis, treatment-related adverse events were defined as those with an attribution score of 3 or higher. To prevent inflation of event counts from recurrent episodes of the same toxicity across cycles, each patient was counted once per adverse event term at the maximum grade observed. Serious adverse events were recorded and reported according to protocol-defined reporting requirements and institutional guidelines.

### Statistical Analysis

Given the rarity of metastatic PPGL and the exploratory nature of this phase II study, all analyses were descriptive. ORR was calculated as the proportion of evaluable patients with a confirmed complete or partial response according to RECIST version 1.1. Time-to-event end points (PFS, OS) were estimated using the Kaplan-Meier method; patients without a documented event were censored on the date of the last adequate disease assessment. The safety population included all patients who received at least one dose of axitinib. The efficacy population included patients who completed at least one post-baseline radiographic assessment. Analyses were performed using SAS version 9.4 (SAS Institute, Cary, NC) or R version 4.3 (R Foundation for Statistical Computing, Vienna, Austria).

### Study Conduct and Enrollment

All patients provided written informed consent and were enrolled by the same investigator (TF) at two different institutions. At the National Cancer Institute, patients were enrolled between September 9, 2014, and May 3, 2015. At Columbia University / New York Presbyterian Hospital, patients were enrolled between January 22, 2019, and February 13, 2023. Two patients enrolled at the National Cancer Institute were unable to transfer care to Columbia University because of financial reasons and continued follow up at the National Cancer Institute. The interval in enrollment between institutions reflected administrative and contractual delays associated with transferring study conduct to Columbia University. The initial data cutoff occurred on May 30, 2024.

## RESULTS

### Patient Characteristics

Nineteen patients were enrolled between December 2013 and May 2015. Seventeen patients received at least one dose of axitinib and constituted the safety and efficacy populations. Two patients discontinued the study before receiving protocol treatment and were therefore not included in the efficacy analyses. Baseline demographic and clinical characteristics are summarized in **Table 1**. The median age was 53 years (range, 18-70 years). All treated patients had histologically confirmed metastatic PPGL with radiographically documented disease progression before enrollment. The study population included patients with both adrenal pheochromocytoma and extra-adrenal paraganglioma, with a high prevalence of hereditary disease. Most patients had received one or more prior locoregional or systemic therapies, including surgery, external beam radiotherapy, high-specific-activity ^131^I-metaiodobenzylguanidine (^131^I-MIBG), peptide receptor radionuclide therapy, cyclophosphamide/vincristine/dacarbazine (CVD) chemotherapy, and temozolomide,

**Table 1.** Baseline Demographic and Clinical Characteristics.

| **Table 1. Baseline Demographic and Clinical Characteristics** |  |
| --- | --- |
| **Characteristic** | **Overall (N = 17)** |
| **Age, years: median (range)** | **53 (18–70)** |
| **Sex: Male (%) / Female (%)** | **14 (82.4) / 3 (17.6)** |
| **Primary Tumor, n (%)** |  |
| Pheochromocytoma | **11 (64.7)** |
| Paraganglioma | **6 (35.3)** |
| **Molecular/hereditary status, n (%)** |  |
| *SDHB* | **8 (47.1)** |
| *NF1* | **1 (5.9)** |
| Beckwith-Wiedemann syndrome | **1 (5.9)** |
| No pathogenic germline variant identified | **7 (41.2)** |
| *Previous therapies | • Surgery<br>• External beam radiotherapy<br>• High-specific-activity <sup>131</sup>I-metaiodobenzylguanidine (<sup>131</sup>I-MIBG)<br>• Peptide receptor radionuclide therapy (PRRT; Lutathera®)<br>• Cyclophosphamide, vincristine, dacarbazine (CVD)<br>• Temozolomide |

\*Patients may have received more than one prior therapy

### Treatment Exposure

All patients initiated axitinib at the protocol-specified starting dose of 5 mg twice daily. Treatment was administered continuously in 28-day cycles, with protocol-defined dose escalation or reduction based on individual tolerability. The median duration of treatment was 9.5 months. Dose modifications were primarily required for hypertension and other expected VEGFR inhibitor-associated toxicities. No patient permanently discontinued axitinib because of treatment-related toxicity, and temporary dose interruptions successfully permitted continuation of therapy following toxicity resolution. Treatment-related toxicities were managed with temporary treatment interruptions, dose reductions, and supportive care. One patient remained on treatment for 70 cycles at the time of data cutoff, demonstrating prolonged clinical benefit.

### Antitumor Activity

Seventeen treated patients were evaluable for efficacy. Six patients achieved a confirmed partial response, corresponding to an objective response rate of 35.3%. The remaining patients had stable disease or progressive disease as their best overall response. No complete responses were observed. The median maximum reduction in RECIST target lesion measurements among responding patients was 34% **(Figure 1)**, and the median duration of response was 7.4 months. For the entire population, median progression-free survival (PFS) was 7.9 months (95% CI, 6.1-16.8 months), and median overall survival (OS) was 29 months. Kaplan–Meier estimates of PFS are shown in **Figure 2**. Durable disease control was observed in a subset of patients, including one patient who remained on treatment for more than 70 treatment cycles. Individual treatment duration and best overall response are illustrated in the swimmer plot **(Figure 3)**, demonstrating sustained clinical benefit in several patients, including ongoing treatment in two patients at the time of data cutoff.

**Figure 1.**
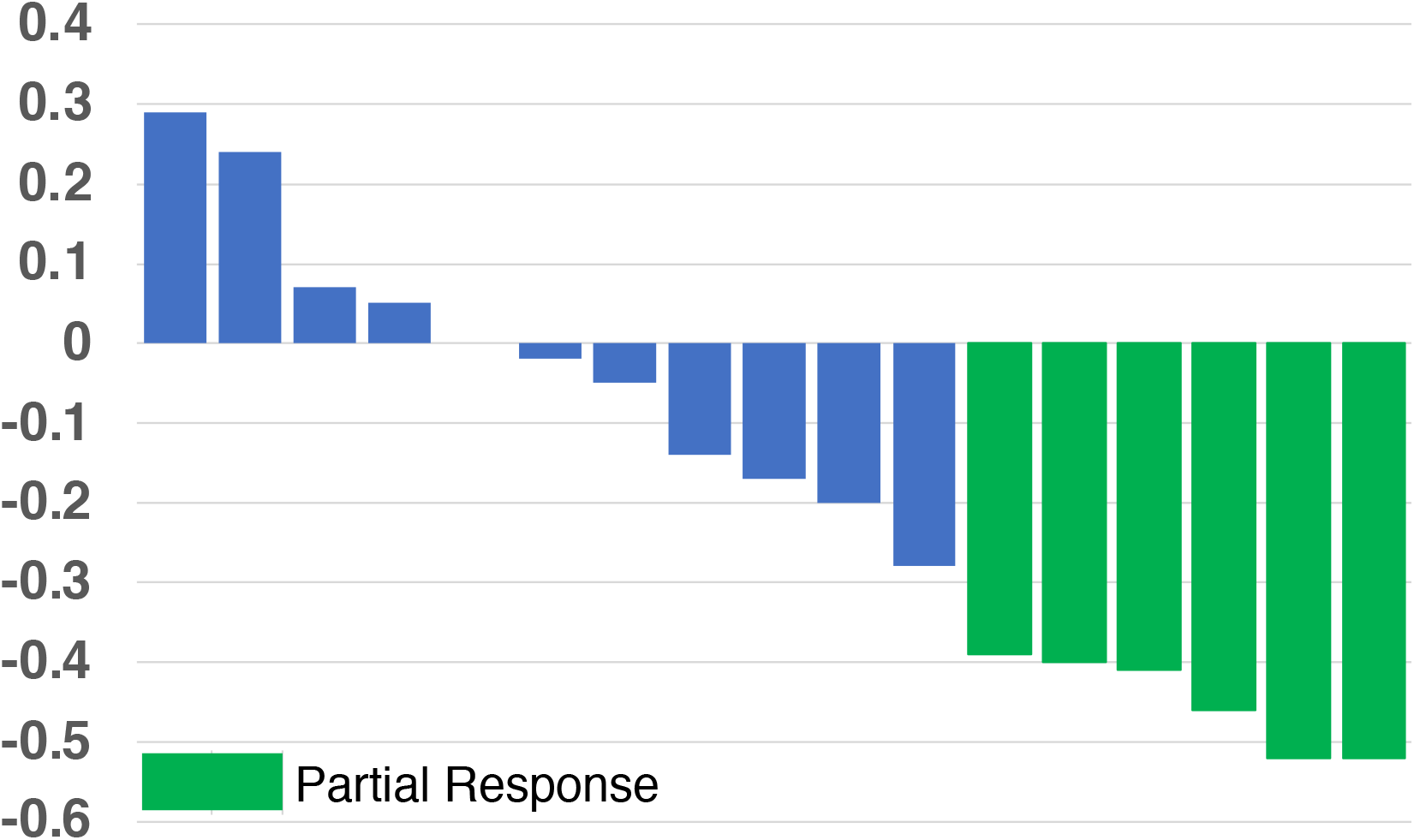
Waterfall plot showing the maximum percentage change from baseline in the sum of RECIST target lesion diameters in response-evaluable patient treated with axitinib.

**Figure 2.**
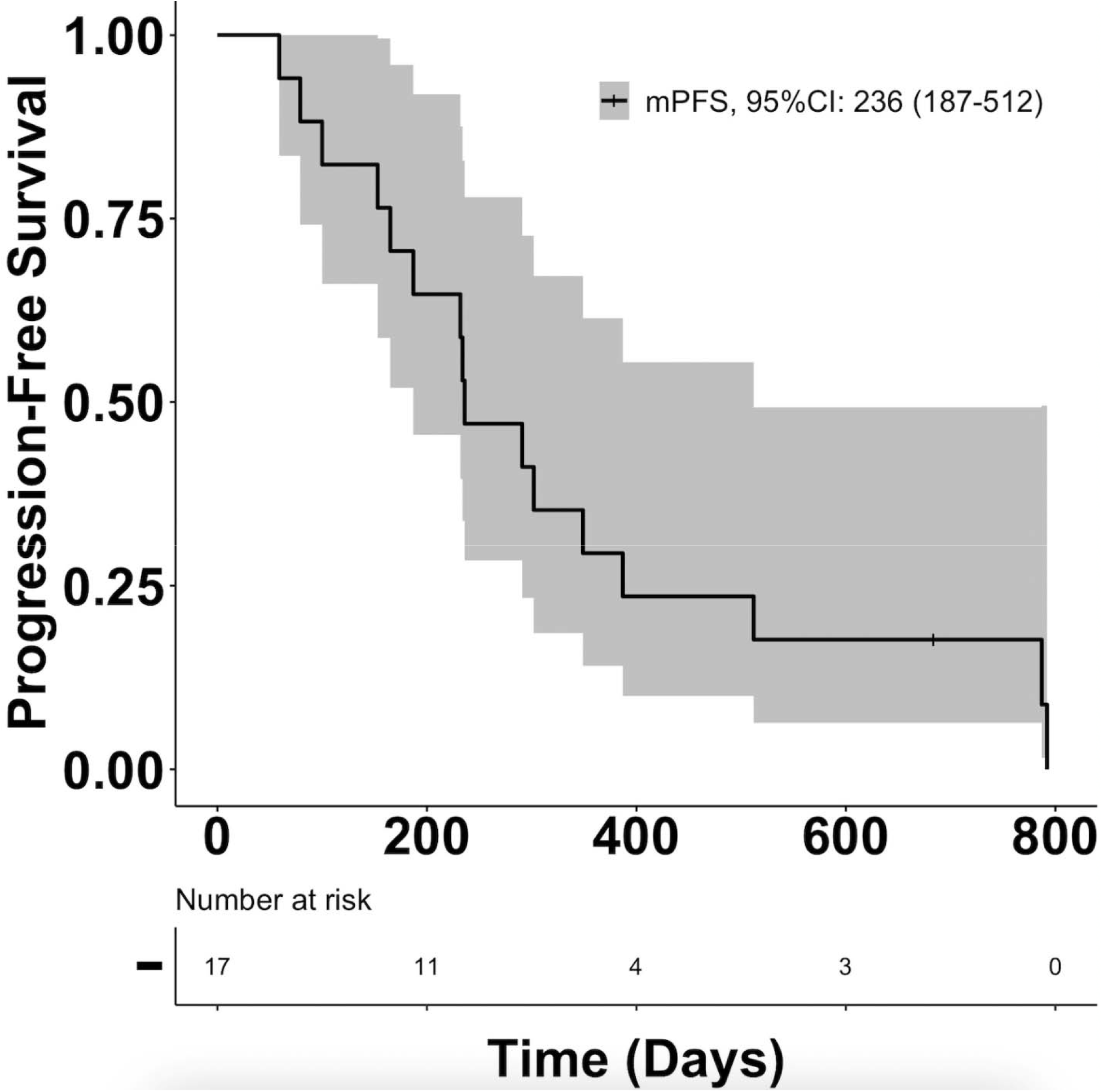
Kaplan-Meier estimate of progression-free survival (PFS) in patients treated with axitinib. PFS was measured from initiation of study treatment until radiographic disease progression or death from any cause. Patients without a PFS event were censored at the date of their last disease assessment. Tick marks indicate censored observations. The shaded area represents the 95% confidence interval. Median PFS was 236 days (95% CI, 187-512 days), corresponding to 7.9 months (95% CI, 6.1-16.8 months). The number of patients at risk is shown below the x-axis.

**Figure 3.**
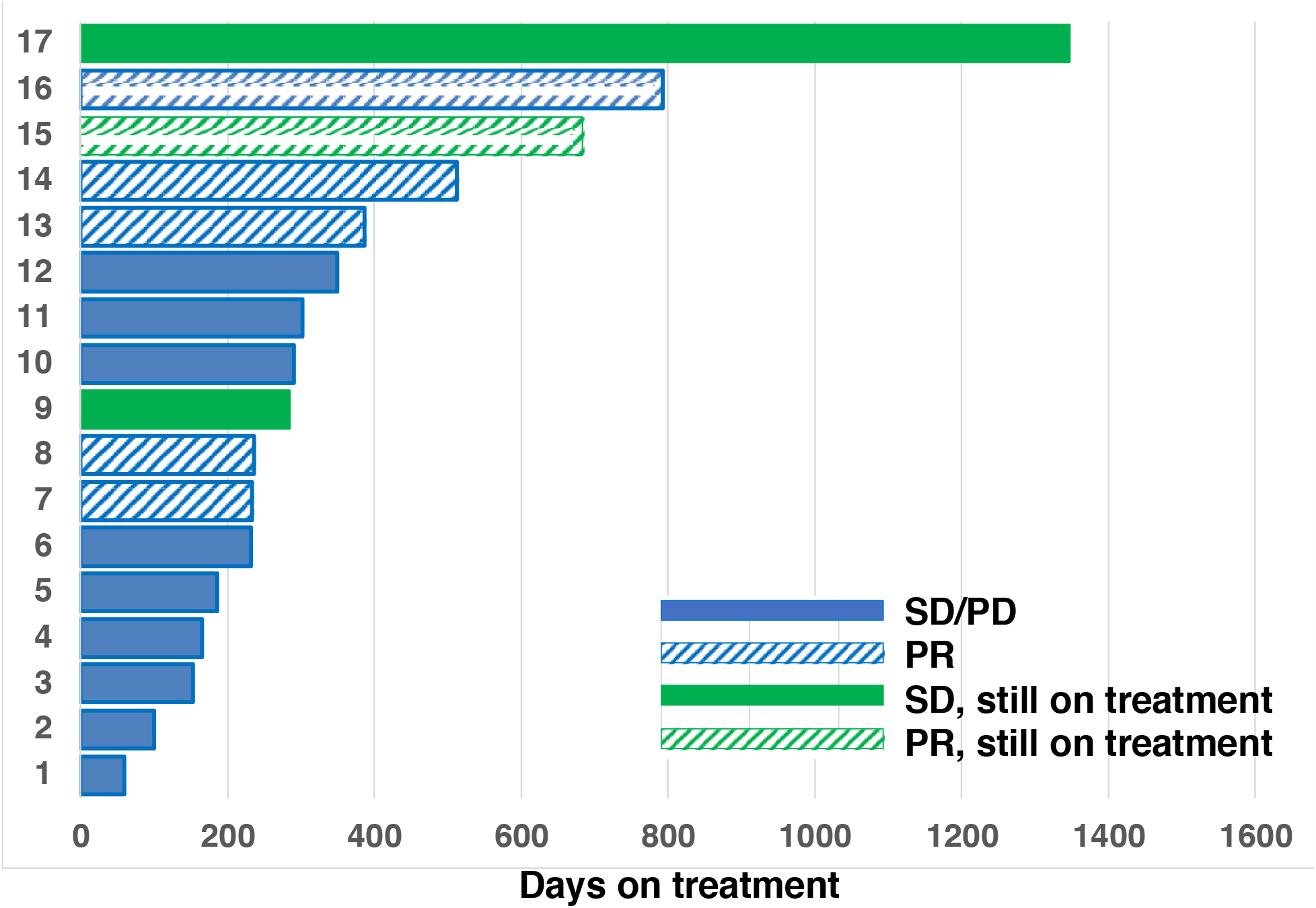
Swimmer plot of treatment duration and best overall response for individual patients treated with axitinib. Each horizontal bar represents one patient and depicts the duration of treatment. Bar patterns indicate best overall response according to RECIST v1.1 (partial response or stable/progressive disease). Solid green bars represent patients with ongoing stable disease, and hatched green bars represent patients with ongoing partial response at the time of data cutoff.

### Safety

Safety analyses included all 17 treated patients, with adverse events summarized according to the highest CTCAE grade reported for each patient for each adverse event. Treatment-related adverse events were consistent with the known safety profile of axitinib **(Table 2)**. Hypertension was the most frequently observed treatment-related adverse event, occurring in 79% of patients, and was the most common toxicity requiring dose modification and antihypertensive therapy. Blood pressure was managed with antihypertensive therapy and protocol-directed dose adjustments, with no permanent treatment discontinuations because of hypertension.

**Table 2:** Treatment-Related Adverse Events in Patients Treated with Axitinib [N = 17].

|  | Grade 1 | Grade 2 | Grade 3 |
| --- | --- | --- | --- |
| Abdominal pain | 5 | 2 |  |
| Anorexia | 1 | 4 |  |
| Anxiety | 1 |  |  |
| Arthralgia | 1 | 2 |  |
| Constipation | 2 |  |  |
| Cough | 3 |  |  |
| Diarrhea | 9 | 2 |  |
| Dizziness | 1 | 1 |  |
| Dry skin | 2 |  |  |
| Dysgeusia | 3 | 2 |  |
| Dysphagia | 1 | 1 |  |
| Dyspnea | 3 |  |  |
| Fatigue | 3 | 9 |  |
| Headache | 4 | 2 |  |
| Hoarseness | 7 |  |  |
| Hypertension | 1 | 4 | 10 |
| Insomnia | 2 |  |  |
| Mucositis oral | 7 | 1 | 2 |
| Nausea | 7 | 1 |  |
| Pain in extremity | 1 | 3 |  |
| Palmar-plantar erythrodysesthesia | 3 | 5 |  |
| Peripheral sensory neuropathy | 2 |  |  |
| Rash acneiform | 3 |  |  |
| Rash maculo-papular | 1 | 1 |  |
| Vomiting | 2 | 3 |  |
| Weight loss |  | 2 |  |
| **Laboratory Adverse Events** |  |  |  |
| ↑ ALT | 2 |  |  |
| Anemia | 2 |  | 1 |
| Hypothyroidism | 1 | 5 |  |
| ↓ Neutrophil count |  | 2 |  |
| ↓ Platelet count | 4 |  |  |
| Proteinuria | 1 | 2 |  |
| ↓ WBC | 4 | 1 |  |

Grade ≥3 treatment-related clinical and laboratory adverse events are summarized in **Table 2**. In addition to hypertension, the most common Grade 3 treatment-related toxicities included anemia, oral mucositis, hypophosphatemia, duodenal hemorrhage, and decreased lymphocyte count. Most remaining treatment-related adverse events were Grade 1 or 2 and were effectively managed with supportive care, temporary treatment interruption, or dose reduction. Serious adverse events were uncommon and occurred in two patients. Reported serious adverse events included Grade 3 anemia (n=1), and Grade 3 vaginal hemorrhage (n=1). All serious adverse events were successfully managed with supportive care and protocol-directed interventions, and none resulted in permanent discontinuation of study treatment. Investigator assessment classified all serious adverse events as unlikely related to axitinib, and no treatment-related deaths occurred.

## DISCUSSION

This prospective phase II study demonstrated that axitinib has clinically meaningful antitumor activity with treatment-related toxicities that were consistent with the known safety profile of VEGF receptor inhibition in other solid tumors, and generally manageable with standard supportive measures and protocol-directed dose modifications. No patient permanently discontinued treatment because of treatment-related toxicity.

The randomized FIRSTMAPPP trial established sunitinib as the first VEGF-targeted therapy to demonstrate a significant PFS benefit in patients with progressive metastatic PPGL, with an ORR of 36.1% in the sunitinib arm and a 12-month PFS rate of 36% (vs. 19% with placebo). The FIRSTMAPPP trial also reported a 50% partial response rate in patients with SDHB mutations, providing exploratory clinical support for the potential sensitivity of pseudohypoxia-associated tumors to VEGF-targeted therapy. [1] Although direct comparisons between studies should be interpreted cautiously because of differences in study design, eligibility criteria, and sample size, the efficacy observed in the present cohort appears generally consistent with that reported for sunitinib as shown in **Table 3**. A meta-analysis of TKIs in metastatic PPGL, encompassing 160 patients across 7 studies, reported a pooled partial response rate of 32%, disease control rate of 86%, and a combined median PFS of 8.9 months. [2] Because this meta-analysis included data from the first nine patients treated in the present axitinib study, these results should not be considered independent validation of the current findings. Together, these data support VEGF pathway inhibition as an active therapeutic strategy in metastatic PPGL and provide a rationale for selective VEGF receptor inhibition with axitinib. [13, 17, 24]

**Table 3:** Selected Efficacy and Safety Outcomes Reported with Sunitinib in FIRSTMAPPP and Axitinib in the Present Study.

|  | **Sunitinib**<br>**[FIRSTMAPPP Study]** | **Axitinib**<br>**[This study]** |
| --- | --- | --- |
| Objective Response Rate | 31% | 35% |
| Median PFS | 8.9 months | 8.1 months |
| Median duration of therapy | 11 months | 9.5 months |
| Patients with Grade $\geq 3$ TRAEs, % | 72% (all-cause) | 59% |
| Most common AEs | Hypertension; Fatigue;<br>Asthenia | Hypertension; Fatigue;<br>Mucositis; Diarrhea |
| Drug discontinuation | 14% | 0% |

Abbreviations: TRAE, treatment related adverse event; PFS, progression-free survival.

An important consideration in conducting trials in PPGL is the management of hypertension and other cardiovascular manifestations related to catecholamine excess, particularly when evaluating VEGF pathway inhibitors that can themselves increase blood pressure. Eligibility criteria in this study were intended to facilitate enrollment of a clinically representative (real-world) population while maintaining appropriate safety safeguards. Not surprisingly, hypertension emerged as the most common treatment-related adverse event but was effectively managed through optimization of antihypertensive therapy and protocol-directed dose modification. No patient required permanent treatment discontinuation because of hypertension or other treatment-related toxicities. This agrees with the low 14% discontinuation rate observed with sunitinib in FIRSTMAPPP and the pooled TKI discontinuation rate of 14.3% reported in a meta-analysis.[1, 2] and compares favorably with the single-arm clinical trials in renal cell carcinoma in which the discontinuation rates of axitinib due to AEs were 19% and 25% and 13.8% in the AXIS trial [22, 25, 26] The ability to manage VEGF-associated toxicities without permanent treatment discontinuation in this cohort is clinically relevant. These findings support the feasibility of enrolling patients with PPGL in appropriately designed clinical trials when close blood pressure monitoring and multidisciplinary management are incorporated into study care.

Beyond hypertension, the overall toxicity profile observed in this study was consistent with the established safety profile of axitinib in other solid tumors. Although grade 3 adverse events occurred in several patients, most resolved with temporary treatment interruption, dose reduction, or standard medical management. The ability to maintain patients on treatment despite these toxicities may be particularly relevant when prolonged therapy is required for disease control in slowly progressive neuroendocrine malignancies.

The treatment landscape for metastatic PPGL has evolved substantially since the initiation of this trial. In addition to CVD chemotherapy and radiopharmaceutical therapies, prospective data now support systemic treatment with VEGF-targeted TKIs and HIF-2α inhibition.[26] [2] Results from our study, together with FIRSTMAPPP and other studies of VEGF-directed TKIs, provide prospective evidence supporting VEGF pathway inhibition in metastatic PPGL. Additionally, in May 2025, belzutifan, a selective HIF-2α inhibitor, received FDA approval for locally advanced, unresectable, or metastatic PPGL based on the LITESPARK-015 trial [14], which demonstrated an ORR of 26%, a median duration of response of 20.4 months, and median PFS of 22.3 months. Notably, belzutifan was associated with a reduction in antihypertensive medication in 32% of patients, a potentially clinically relevant effect in this population with catecholamine-induced hypertension.[13]. The NATALIE trial [15] also reported activity with cabozantinib. In the absence of prospective head-to-head trials, the optimal sequencing of these therapies remains undefined. Treatment selection should therefore be individualized according to disease aggressiveness, molecular characteristics, functional status, prior therapies, radiopharmaceutical eligibility, comorbidities, anticipated toxicities, drug availability, and patient preference. Access and cost are also important considerations, particularly in resource-limited settings, and may substantially influence treatment selection and sequencing. However, formal comparative effectiveness and pharmacoeconomic analyses are needed to define the relative value of available therapies.

Several limitations should be acknowledged. First, this study was conducted in a small cohort of patients, reflecting the rarity of metastatic PPGL and the inherent challenges of conducting prospective clinical trials in uncommon malignancies. Second, the single-arm design without a concurrent control group precludes direct comparisons with other systemic therapies and limits definitive assessment of comparative efficacy. Finally, premature study termination by the sponsor limited patient accrual and may have reduced the precision of efficacy estimates and the ability to fully characterize long-term outcomes.

Despite these limitations, this prospective phase II study demonstrated clinically meaningful antitumor activity with axitinib and a manageable toxicity profile. Together with the FIRSTMAPPP trial [2] and other prospective studies, these findings support VEGF pathway inhibition as a rational therapeutic strategy for metastatic PPGL and identify axitinib as a potential treatment option.

## Data Availability

All data produced in the present study are available from the corresponding author upon reasonable request, subject to applicable patient privacy and institutional requirements

